# Quantitative measurement of infectious virus in SARS-CoV-2 Alpha, Delta and Epsilon variants reveals higher infectivity (viral titer:RNA ratio) in clinical samples containing the Delta and Epsilon variants

**DOI:** 10.1101/2021.09.07.21263229

**Authors:** Hannah W. Despres, Margaret G. Mills, David J. Shirley, Madaline M. Schmidt, Meei-Li Huang, Keith R. Jerome, Alexander L. Greninger, Emily A. Bruce

## Abstract

**Background:** Novel SARS-CoV-2 Variants of Concern (VoC) pose a challenge to controlling the COVID-19 pandemic. Previous studies indicate that clinical samples collected from individuals infected with the Delta variant may contain higher levels of RNA than previous variants, but the relationship between viral RNA and infectious virus for individual variants is unknown.

**Methods:** We measured infectious viral titer (using a micro-focus forming assay) as well as total and subgenomic viral RNA levels (using RT-PCR) in a set of 165 clinical samples containing SARS-CoV-2 Alpha, Delta and Epsilon variants that were processed within two days of collection from the patient.

**Results:** We observed a high degree of variation in the relationship between viral titers and RNA levels. Despite the variability we observed for individual samples the overall infectivity differed among the three variants. Both Delta and Epsilon had significantly higher infectivity than Alpha, as measured by the number of infectious units per quantity of viral E gene RNA (6 and 4 times as much, p=0.0002 and 0.009 respectively) or subgenomic E RNA (11 and 7 times as much, p<0.0001 and 0.006 respectively).

**Conclusion:** In addition to higher viral RNA levels reported for the Delta variant, the infectivity (amount of replication competent virus per viral genome copy) may also be increased compared to Alpha. Measuring the relationship between live virus and viral RNA is an important step in assessing the infectivity of novel SARS-CoV-2 variants. An increase in the infectivity of the Delta variant may further explain increased spread and suggests a need for increased measures to prevent viral transmission.

**SIGNIFICANCE STATEMENT:** Current and future SARS-CoV-2 variants threaten our ability to control the COVID-19 pandemic. Variants with increased transmission, higher viral loads, or greater immune evasion are of particular concern. Viral loads are currently measured by the amount of viral RNA in a clinical sample rather than the amount of infectious virus. We measured both RNA and infectious virus levels directly in a set of 165 clinical specimens from Alpha, Epsilon or Delta variants. Our data shows that Delta is more infectious compared to Alpha, with ∼ six times as much infectious virus for the same amount of RNA. This increase in infectivity suggests increased measures (vaccination, masking, distancing, ventilation) are needed to control Delta compared to Alpha.

## MAIN TEXT

Severe acute respiratory syndrome coronavirus 2 (SARS-CoV-2), the virus which causes COVID-19, is responsible for 214,514,637 cases and 4,472,553 deaths as of August, 2021^1^. Despite the rapid development of multiple effective vaccines, the emergence of genetically distinct viral lineages pose challenges to the ability to control the ongoing COVID-19 pandemic^2^. Changes in the viral genome as a result of mutations introduced by the viral polymerase can result in viral variants that have increased levels of transmission, replication and/or impact the effectiveness of current vaccines. These properties are assessed for variants detected by next generation sequencing and the World Health Organization classifies them as Variants of Interest (VOI) or Variants of Concern (VOC) depending on these factors^3^. In the fall of 2020, the Alpha (B.1.1.7) variant emerged in the U.K. and was associated with increased transmission and spread before being designated a VOC in December, 2020^4,5,6,7,8^. By late November of 2020, the Epsilon (B.1.429/B.1.427) lineage emerged in the US state of California^9^ with rapid spread and signs of partial immune evasion, which resulted in its classification as a VOI by the WHO in March of 2021, before being downgraded in the summer of 2021 as Epsilon’s circulation was replaced by a newer variant^8,10,11^. Spread of the Delta (B.1.617.2) variant, first detected in India in the spring of 2021, appears to have dominated both the Alpha and Epsilon variants and now accounts for approximately 90% of viral sequences sequenced globally^12^. This increase in global spread, combined with a possible increase in disease severity compared to Alpha led to Delta being classified as a VOI in April 2021, and a VOC the following month^8,13^. The rise in transmission of the Delta variant appears to be at least in part due to increased viral fitness conferred by mutations in the furin cleavage site that render the virus able to more efficiently enter cells^14,15^. Furthermore, viral RNA levels in samples from people infected with Delta are reported to be higher than those of previous variants with an increased duration of viral shedding^16,17,18,19^. Of particular concern, the levels of RNA in upper respiratory samples from infected individuals are reported to be similar among vaccinated and unvaccinated individuals^18,20^.

While increasing genomic surveillance has improved the ability to detect viral lineages in near real-time across many geographic areas, further investigation into the biological characteristics of these variants is required to better understand which mutations are responsible for observed increases in viral transmission. One difficulty of interpreting viral infectivity lies in the widespread use of viral RNA levels (measured by reverse transcriptase quantitative polymerase chain reaction [RT-PCR] in clinical samples used for SARS-CoV-2 diagnosis) as measurements of viral load. While it is logical to assume that there is a relationship between RNA and infectious viral levels, it is unlikely this is a fixed ratio in all scenarios. The genome to plaque forming units (PFU) ratio for SARS-CoV-2 has been reported to be in the range 10^3^-10^6^:1, while the original SARS is thought to be comparatively more infectious per particle, with a ratio of 360 genomes to each PFU^21,22^. Quantitative measurement of replication competent virus by plaque or focus assay would improve the ability to determine and interpret infectious viral loads for current and future variants, including VOI and VOC.

To measure the infectious viral titers of SARS-CoV-2 clinical specimens, and the relationship between replication competent virus and viral RNA, we performed viral focus assays and RT-qPCR on a set of 165 SARS-CoV-2 positive samples belonging to the Alpha, Epsilon or Delta variants. To minimize variation resulting from sample handling, we exclusively used samples that had been frozen within two days of sample collection from the patient. Once collected, clinical samples are routinely stored and transported at 4°C and our previous work demonstrated that infectious virus and RNA are stable at both 4°C and room temperature for over four days^23^. While limited residual material in clinical samples makes performing traditional plaque assays technically challenging, we have successfully used a focus forming assay performed in 96-well plates to measure infectious viral titer of volumes as small as 100-200 µl^23,24^. This approach enables us to measure individual infectious viral units directly from residual clinical samples, which to our knowledge are the first such measurements reported.

As expected, we observed a positive correlation between the amount of viral E gene RNA and the level of infectious virus for all three variants (Fig. 1, Table S1). This was true for both total levels of RNA and E subgenomic RNA, as measured by the cycle threshold (C_T_) in which a sample is detected by RT-PCR, where a lower C_T_ indicates more RNA. Surprisingly however, there was a high level of variability in the ratio of RNA to virus, particularly in Delta, as some samples with similar C_T_ values had viral titers over three logs (1,000-fold) apart. Despite the variability in RNA to infectious virus ratios for individual samples, we observed different trends of overall infectivity between the three variants we studied. Both Epsilon and Delta had significantly higher infectivity than Alpha, as measured by the number of infectious units per quantity of viral RNA (Fig. 1A). We generated lines of best fit for each variant using linear regression on log transformed data and based on this model our data suggests that the Delta and Epsilon SARS-CoV-2 variants have 5.9 and 4.3 times as much infectious virus than Alpha, for samples with the same amount of total viral RNA (p=0.0002 and 0.009 respectively). We observed a similar trend when comparing infectious virus to subgenomic E RNA levels (11.3-fold, p<0.0001; 7.3-fold, p=0.006) (Fig. 1B). While subgenomic RNA has been proposed as a specific marker of infectious virus^25,26^, we observed a broad range in the ratio of sgE RNA to infectious virus for our sample set (similar to that seen for total RNA), in line with previous reports suggesting that subgenomic RNA levels are not necessarily indicative of actively replicating virus^27,28,29^.

**Figure 1.**
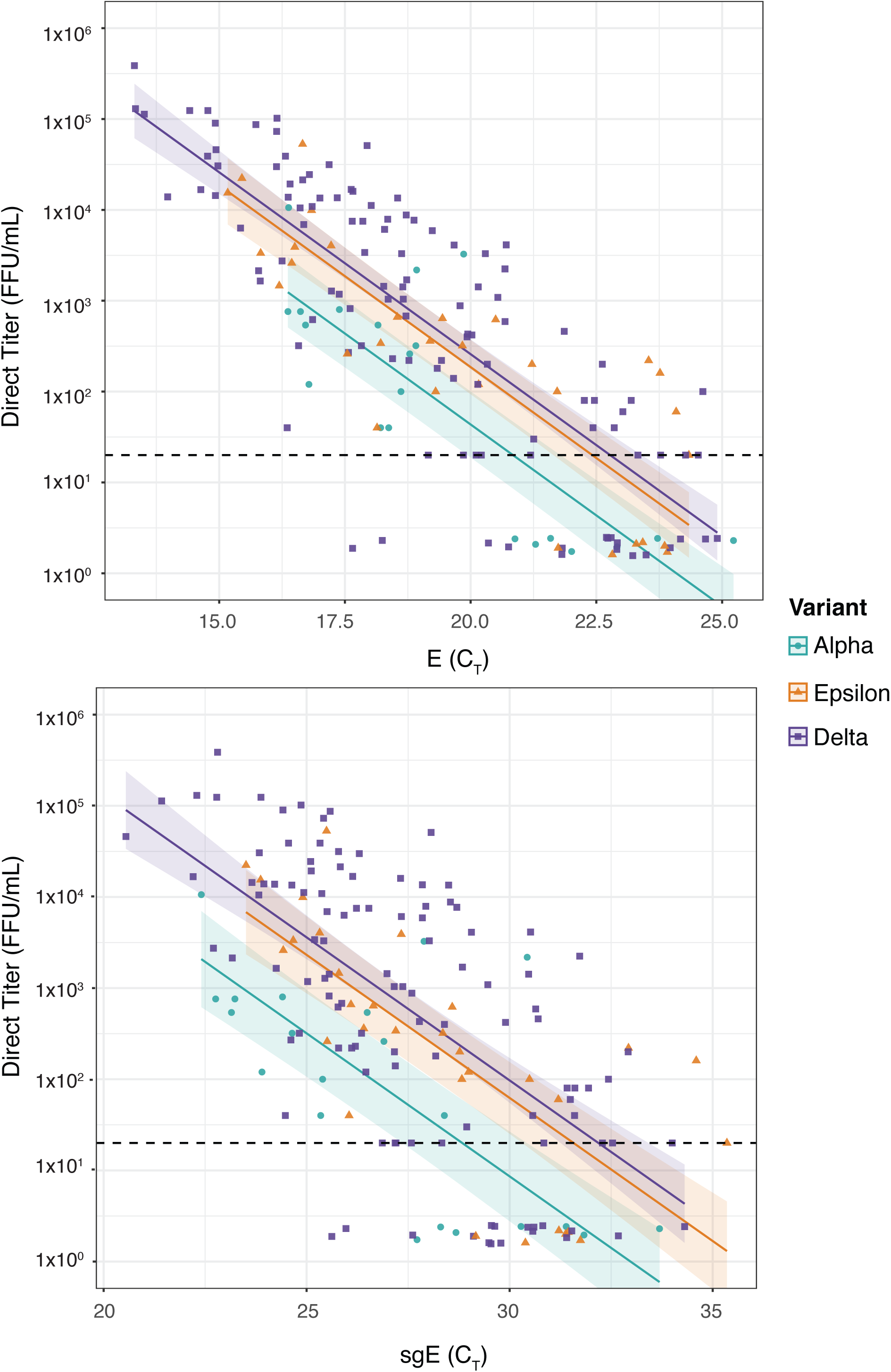
Epsilon and Delta SARS-CoV-2 variants have a higher ratio of infectious virus to RNA than the Alpha variant in clinical specimens. A set of 165 clinical swab samples from individuals infected with the Alpha (teal circles; n=21), Epsilon (orange triangles; n=31) or Delta (purple squares; n=55+58) SARS-CoV-2 variants of concern was used to determine the relationship between viral titer and viral RNA levels for each sample. **A)** For each clinical sample total E RNA (RT-PCR C_T_ detected by WHO-E primer/probe set^38^) and **B)** subgenomic E RNA (RT-PCR C_T_ detected by the Mills-sgE primer set we previously developed^23^) is plotted on the X axis. Viral titer for each sample (FFU/mL measured by viral focus assay) is plotted on the Y axis. The limit of detection for infectious titer is 20 FFU/mL and is indicated with a dashed line. Samples for which we could not measure a viral titer were randomly assigned a value between 1.5-2.5 for display purposes. Lines of best fit and 95% confidence intervals were generated by linear regression on log transformed data, with un-detected viral titers assigned fixed values of one tenth the Limit of Detection, i.e. 2 FFU.

A limitation of this study is the lack of access to metadata for the clinical specimens (including C_T_ from targets other than E/sgE from diagnostic platforms), which may obscure differences in age, pre-existing conditions, days from exposure or symptom onset, and vaccination status. Furthermore, due to the timing of variant spread across the Pacific Northwest (and USA in general), the three variants for which we present data were not all circulating at the same time, and thus we cannot compare samples for all three variants collected simultaneously. We have attempted to address this by analyzing Alpha and Epsilon samples that were collected at the same time, and thus are drawn from a population that would be expected to have similar vaccination completion rates. In addition, we analyzed data for Delta samples collected on two different days (n=55 and 59) and did not observe a statistically significant batch effect. Recent work from the Klein, Pekosz and Mostofa groups indicates that infection with the Delta variant is associated with a higher likelihood of infectious virus isolation compared to Alpha, for both vaccinated and unvaccinated individuals^18^, unlike previous work indicating that the likelihood of infectious virus isolation was reduced in vaccinated individuals infected with Alpha^30^. Although we do not have access to information on vaccination status for our dataset, our data supports the finding that higher infectious viral loads are present in Delta vs Alpha samples.

The data presented here suggests that measuring ‘viral load’ strictly by the amount of RNA present in a clinical sample may have limitations. Variability in sample handling, vaccination status and days post symptom onset are all potential contributors to the variation we observed in genome:focus forming unit (FFU) ratios amongst clinical specimens. However, it is well established that the ‘particle:pfu’ ratio for viruses can vary between viral strains and is influenced by the cell type or organism the viral sample has been grown in^22,31,32^. Our data indicate that the infectivity of specific SARS-CoV-2 variants may also vary. We propose that the relationship between viral RNA and infectious virus should be investigated for future variants rather than relying on RNA C_T_ as the sole measurement of viral load. The finding that the Delta variant shows increased infectivity compared to Alpha is in line with the increased transmission and spread observed for this variant^17^, as well as reports indicating live virus is more likely to be isolated from Delta clinical specimens compared to Alpha^18^. We observed similar infectivity profiles for Delta and Epsilon; however we saw both increased infectivity and an increase in overall RNA levels for Delta samples. The dual advantage of higher replication (ie, RNA levels) and higher specific particle infectivity (FFU:genome) may provide a possible explanation for Delta overtaking Epsilon in the United States during the summer of 2021. The increased infectivity we observe in Delta clinical samples underscores the need for increased measures to prevent transmission to those who remain vulnerable, such as widespread vaccination, masking, distancing, and improved ventilation.

## METHODS

### Selection of samples and typing of SARS-CoV-2 variants using RT-ddPCR and NGS surveillance

Clinical specimens (mostly nasal swabs) that had newly been identified as positive for SARS-CoV-2 by any of the clinical testing platforms in use at the University of Washington Virology Laboratory (UWVL) were selected on three different days: March 25, 2021, August 3, 2021 and August 26, 2021. Aliquots from each sample were taken within two days of sample collection, de-identified, and frozen at -80 C. The use of deidentified positive specimens for the above studies was approved by the University of Washington Institutional Review Board (STUDY00010205). Mutations indicating variants of concern were identified in clinical specimens using an RT-ddPCR assay similar to that we have previously described in Perchetti et al 2021^33^.The assay screens for mutations in the region of the Spike gene encoding the Receptor Binding Domain: K417N/T; L452R; E484K/Q; and N501Y. The results were interpreted based on NGS surveillance done by UWVL in the period of each sample collection. In the week of the March collection, surveillance sequencing identified 31.5% Alpha, 29.5% Epsilon, and 0.1% Delta; in the weeks of the August collection, samples were 0.4-0.6% Alpha, 0% Epsilon, and 98.1-98.5% Delta (https://depts.washington.edu/labmed/covid19/#sequencing-information). For the March collection date, Alpha was indicated by 417K, 452L, 484E, and 501Y; Epsilon was indicated by 417K, 452R, 484E, and 501N. For the August collection dates, Delta was indicated by 417K/N, 452R, 484E/Q, and 501N. Samples not matching these three variant designations were identified in both collections but we did not identify enough of any single additional variant/Pango lineage to be useful for statistical comparison.

### RNA extractions

Total nucleic acid (TNA) in all clinical samples were extracted with an automated guanidinium lysis/magnetic silica bead absorption method using MagNA Pure 96 instrument and DNA & Viral NA Small Volume kit (Roche, Cat. # 05 467 497 001) according to manufacturer instructions. All extractions used 200 µl of input volume and were eluted into 50 µl.

### RT-PCR

Specific viral RNA was reverse transcribed into cDNA and then amplified in real-time PCR reactions using AgPath-ID One Step RT-PCR kit (Life Technologies, ThermoFisher, Cat. #4387424M), using 5 µl of extracted TNA per 25 µl reaction. RT-PCR reactions used one of two sets of primers/probes: 1) WHO-E, using the E_Sarbeco-F/R/P set^34^; 2) Mills-sgE, using sgLeader-F2, sgE-R, and sgE-P^23^. RT-PCR was performed on an ABI 7500 real-time PCR system as previously described^35^.

### Focus forming assay

SARS-CoV-2 viral isolations were conducted at the UVM BSL-3 facility under an approved Institutional Biosafety protocol. Viral titer was determined by focus forming assay in a 96 well plate format. Serial ten-fold dilutions of clinical sample were used to inoculate Vero E6-TMPRSS2 cell monolayers (60,000 cells/well seeded one day prior) in 96□well white polystyrene microplates (Falcon, Cat. # 08 771 26). 50 µL of each virus dilution was inoculated onto the cells and incubated at 37°C in a 5% CO_2_ incubator for 60 min, after which the wells were overlaid with 1.2% methylcellulose (Acros cat. #332620010) in DMEM (Gibco, cat. #11965 092) and incubated at 37°C in a 5% CO_2_ incubator for 24 h. All dilutions were plated in duplicate with the exception of the neat clinical sample for which we only had sufficient material to plate one well per sample. Infected cells were fixed in 4% formaldehyde in 3 × PBS. Cells were permeabilized with 0.1% 100X Triton in 1× PBS for 15 min and then incubated with a primary, cross-reactive rabbit anti-SARS-CoV N monoclonal antibody (Sinobiological, distributed by Thermo Fisher, Cat. #40143-R001 at a dilution of 1:20,000) followed by a peroxidase-labelled goat anti-rabbit antibody (SeraCare, Milford, MA, USA, Cat. #5220-0336 diluted to 1:4,000) and then the peroxidase substrate (SeraCare, Cat. #5510-0030). Plates were imaged on a BioTek ImmunoSpot S6 MACRO Analyzer and foci were counted using an automated virus plaque counter as previously described^36^ and then manually corrected.

### Statistical Analysis

Data were analysed using R^37^. Titers were log transformed, so care was required for the substantial fraction of samples that had titres below the limit of detection (LoD), and several different approaches were considered: removal, assignment to the LoD (20), and assignment of sub-LoD values (LoD/10=2, LoD/100=0.2). Linear regression was employed to model the relationship between titer and C_T_(E) or C_T_(sgE). Interaction terms between the C_T_ values and variant were not significant and were dropped in favor of a simpler variant intercept only model, with all variants sharing a common slope. Models where titers below the LoD were removed remained significant, and had similar effect sizes to the one-tenth LoD approach which was considered optimal. With below-LoD titers removed entirely, the C_T_(E) fold change for Delta vs Alpha was 5.8, while set to LoD (20), LoD/10 (2), and LoD/100 (0.2) before log transform respectively, fold changes were 4.1, 5.9, and 8.4, indicating that setting these values to LoD/10 prior to log transform fits well with the trends observed in the above-LoD values, and does not exert undue influence. A quasi-poisson regression using non-transformed 0 values was also considered and produced comparable estimates of a 6.4-fold change.

## Data Availability

The data supporting the findings of this study are available within the manuscript and supplementary figures. R code is available upon reasonable request.

## ACKNOWLEDGEMENTS

We thank Hannah Kubinski and Joyce Oetjen for technical assistance and Deborah Donnell, Dimitry Krementsov and Beth Kirkpatrick for helpful discussion. This work was supported by the Health and Environmental Sciences Institute (HESI) and NIH P30GM118228-04 (to E.A.B.)

## REFERENCES

1. Covidtracker - Covid-19 Coronavirus Tracker. Accessed August 26, 2021. https://www.covidtracker.com/

2. Callaway E. Delta coronavirus variant: scientists brace for impact. Nature. 2021;595(7865):17–18. doi:10.1038/d41586-021-01696-3

3. The effects of virus variants on COVID-19 vaccines. Accessed September 7, 2021. https://www.who.int/news-room/feature-stories/detail/the-effects-of-virus-variants-on-covid-19-vaccines

4. Davies NG, Abbott S, Barnard RC, et al. Estimated transmissibility and impact of SARS-CoV-2 lineage B.1.1.7 in England. Science. 2021;372(6538). doi:10.1126/science.abg3055

5. Thorne LG, Bouhaddou M, Reuschl A-K, et al. Evolution of Enhanced Innate Immune Evasion by the SARS-CoV-2 B.1.1.7 UK Variant. Microbiology; 2021. doi:10.1101/2021.06.06.446826

6. Parker MD, Lindsey BB, Shah DR, et al. Altered Subgenomic RNA Expression in SARS-CoV-2 B.1.1.7 Infections.; 2021:2021.03.02.433156. doi:10.1101/2021.03.02.433156

7. Brown JC, Goldhill DH, Zhou J, et al. Increased Transmission of SARS-CoV-2 Lineage B.1.1.7 (VOC 2020212/01) Is Not Accounted for by a Replicative Advantage in Primary Airway Cells or Antibody Escape.; 2021:2021.02.24.432576. doi:10.1101/2021.02.24.432576

8. Tracking SARS-CoV-2 variants. Accessed September 7, 2021. https://www.who.int/emergencies/emergency-health-kits/trauma-emergency-surgery-kit-who-tesk-2019/tracking-SARS-CoV-2-variants

9. COVID-19 Variant First Found in Other Countries and States Now Seen More Frequently in California. Accessed August 26, 2021. https://www.cdph.ca.gov/Programs/OPA/Pages/NR21-020.aspx

10. Zhang W, Davis BD, Chen SS, Sincuir Martinez JM, Plummer JT, Vail E. Emergence of a Novel SARS-CoV-2 Variant in Southern California. JAMA. 2021;325(13):1324–1326. doi:10.1001/jama.2021.1612

11. McCallum M, Bassi J, Marco AD, et al. SARS-CoV-2 Immune Evasion by Variant B.1.427/B.1.429.; 2021:2021.03.31.437925. doi:10.1101/2021.03.31.437925

12. GISAID - hCov19 Variants. Accessed August 27, 2021. https://www.gisaid.org/hcov19-variants/

13. Hospital admission and emergency care attendance risk for SARS-CoV-2 delta (B.1.617.2) compared with alpha (B.1.1.7) variants of concern: a cohort study - ScienceDirect. Accessed September 2, 2021. https://www.sciencedirect.com/science/article/pii/S1473309921004758?via%3Dihub

14. Peacock TP, Sheppard CM, Brown JC, et al. The SARS-CoV-2 Variants Associated with Infections in India, B.1.617, Show Enhanced Spike Cleavage by Furin.; 2021:2021.05.28.446163. doi:10.1101/2021.05.28.446163

15. Lubinski B, Frazier LE, Phan MVT, et al. Spike Protein Cleavage-Activation Mediated by the SARS-CoV-2 P681R Mutation: A Case-Study from Its First Appearance in Variant of Interest (VOI) A.23.1 Identified in Uganda.; 2021:2021.06.30.450632. doi:10.1101/2021.06.30.450632

16. Ong SWX, Chiew CJ, Ang LW, et al. Clinical and virological features of SARS-CoV-2 variants of concern: a retrospective cohort study comparing B.1.1.7 (Alpha), B.1.315 (Beta), and B.1.617.2 (Delta). Clinical Infectious Diseases. 2021;(ciab721). doi:10.1093/cid/ciab721

17. Viral infection and transmission in a large well-traced outbreak caused by the Delta SARS-CoV-2 variant - SARS-CoV-2 coronavirus / nCoV-2019 Genomic Epidemiology. Virological. Published July 7, 2021. Accessed August 26, 2021. https://virological.org/t/viral-infection-and-transmission-in-a-large-well-traced-outbreak-caused-by-the-delta-sars-cov-2-variant/724

18. Luo CH, Morris CP, Sachithanandham J, et al. Infection with the SARS-CoV-2 Delta Variant Is Associated with Higher Infectious Virus Loads Compared to the Alpha Variant in Both Unvaccinated and Vaccinated Individuals. Infectious Diseases (except HIV/AIDS); 2021. doi:10.1101/2021.08.15.21262077

19. Teyssou E, Delagrèverie H, Visseaux B, et al. The Delta SARS-CoV-2 variant has a higher viral load than the Beta and the historical variants in nasopharyngeal samples from newly diagnosed COVID-19. J Infect. Published online August 19, 2021. doi:10.1016/j.jinf.2021.08.027

20. Kissler SM, Fauver JR, Mack C, et al. Viral Dynamics of SARS-CoV-2 Variants in Vaccinated and Unvaccinated Individuals.; 2021:2021.02.16.21251535. doi:10.1101/2021.02.16.21251535

21. Vicenzi E, Canducci F, Pinna D, et al. Coronaviridae and SARS-associated Coronavirus Strain HSR1. Emerg Infect Dis. 2004;10(3):413–418. doi:10.3201/eid1003.030683

22. Klimstra WB, Tilston-Lunel NL, Nambulli S, et al. SARS-CoV-2 growth, furin-cleavage-site adaptation and neutralization using serum from acutely infected hospitalized COVID-19 patients. J Gen Virol. 2020;101(11):1156–1169. doi:10.1099/jgv.0.001481

23. Bruce EA, Mills MG, Sampoleo R, et al. Predicting Infectivity: Comparing Four PCR-Based Assays to Detect Culturable SARS-CoV-2 in Clinical Samples.; 2021:2021.07.14.21260544. doi:10.1101/2021.07.14.21260544

24. Graham NR, Whitaker AN, Strother CA, et al. Kinetics and Isotype Assessment of Antibodies Targeting the Spike Protein Receptor Binding Domain of SARS-CoV-2 In COVID-19 Patients as a Function of Age and Biological Sex.; 2020:2020.07.15.20154443. doi:10.1101/2020.07.15.20154443

25. Wong CH, Ngan CY, Goldfeder R, et al. Subgenomic RNAs as molecular indicators of asymptomatic SARS-CoV-2 infectio. Published online 2021. doi:10.1101/2021.02.06.430041

26. Zollo M, Ferrucci V, Izzo B, et al. SARS-CoV-2 Subgenomic N (sgN) Transcripts in Oro-Nasopharyngeal Swabs Correlate with the Highest Viral Load, as Evaluated by Five Different Molecular Methods. Diagnostics. 2021;11(2):288. doi:10.3390/diagnostics11020288

27. Kampen JJA van, Vijver DAMC van de, Fraaij PLA, et al. Shedding of Infectious Virus in Hospitalized Patients with Coronavirus Disease-2019 (COVID-19): Duration and Key Determinants.; 2020:2020.06.08.20125310. doi:10.1101/2020.06.08.20125310

28. Perera RAPM, Tso E, Tsang OTY, et al. SARS-CoV-2 Virus Culture and Subgenomic RNA for Respiratory Specimens from Patients with Mild Coronavirus Disease - Volume 26, Number 11—November 2020 - Emerging Infectious Diseases journal - CDC. doi:10.3201/eid2611.203219

29. Verma R, Kim E, Martínez-Colón GJ, et al. SARS-CoV-2 Subgenomic RNA Kinetics in Longitudinal Clinical Samples. Open Forum Infectious Diseases. 2021;8(7). doi:10.1093/ofid/ofab310

30. Ke R, Martinez PP, Smith RL, et al. Longitudinal Analysis of SARS-CoV-2 Vaccine Breakthrough Infections Reveal Limited Infectious Virus Shedding and Restricted Tissue Distribution. Infectious Diseases (except HIV/AIDS); 2021. doi:10.1101/2021.08.30.21262701

31. McCormick W, Mermel LA. The basic reproductive number and particle-to-plaque ratio: comparison of these two parameters of viral infectivity. Virology Journal. 2021;18(1):92. doi:10.1186/s12985-021-01566-4

32. Alfson KJ, Avena LE, Beadles MW, et al. Particle-to-PFU Ratio of Ebola Virus Influences Disease Course and Survival in Cynomolgus Macaques. Journal of Virology. 89(13):6773–6781. doi:10.1128/JVI.00649-15

33. Perchetti GA, Zhu H, Mills MG, et al. Specific allelic discrimination of N501Y and other SARS-CoV-2 mutations by ddPCR detects B.1.1.7 lineage in Washington State. Journal of Medical Virology. 2021;93(10):5931–5941. doi:10.1002/jmv.27155

34. Wölfel R, Corman VM, Guggemos W, et al. Virological assessment of hospitalized patients with COVID-2019. Nature. 2020;581(7809):465–469. doi:10.1038/s41586-020-2196-x

35. Nalla AK, Casto AM, Huang M-LW, et al. Comparative Performance of SARS-CoV-2 Detection Assays Using Seven Different Primer-Probe Sets and One Assay Kit. Journal of Clinical Microbiology. 58(6):e00557–20. doi:10.1128/JCM.00557-20

36. Katzelnick LC, Escoto AC, McElvany BD, et al. Viridot: An automated virus plaque (immunofocus) counter for the measurement of serological neutralizing responses with application to dengue virus. PLOS Neglected Tropical Diseases. 2018;12(10):e0006862. doi:10.1371/journal.pntd.0006862

37. R: The R Project for Statistical Computing. Accessed September 2, 2021. https://www.r-project.org/

38. Corman V, Bleicker T, Brünink S, et al. Diagnostic detection of 2019-nCoV by real-time RT-PCR. Published online 2020:13.

